# Differential Tractography: A Biomarker for Neuronal Function in Neurodegenerative Disease

**DOI:** 10.1101/2024.08.25.24312255

**Authors:** Connor J. Lewis, Zeynep Vardar, Anna Luisa Kühn, Jean M. Johnston, Precilla D’Souza, William A. Gahl, Mohammed Salman Shazeeb, Cynthia J. Tifft, Maria T. Acosta

## Abstract

GM1 gangliosidosis is an ultra-rare inherited neurodegenerative lysosomal storage disorder caused by biallelic mutations in the *GLB1* gene. GM1 is uniformly fatal and has no approved therapies, although clinical trials investigating gene therapy as a potential treatment for this condition are underway. Novel outcome measures or biomarkers demonstrating the longitudinal effects of GM1 and potential recovery due to therapeutic intervention are urgently needed to establish efficacy of potential therapeutics. One promising tool is differential tractography, a novel imaging modality utilizing serial diffusion weighted imaging (DWI) to quantify longitudinal changes in white matter microstructure. In this study, we present the novel use of differential tractography in quantifying the progression of GM1 alongside age-matched neurotypical controls. We analyzed 113 DWI scans from 16 GM1 patients and 32 age-matched neurotypical controls to investigate longitudinal changes in white matter pathology. GM1 patients showed white matter degradation evident by both the number and size of fiber tract loss. In contrast, neurotypical controls showed longitudinal white matter improvements as evident by both the number and size of fiber tract growth. We also corroborated these findings by documenting significant correlations between cognitive global impression (CGI) scores of clinical presentations and our differential tractography derived metrics in our GM1 cohort. Specifically, GM1 patients who lost more neuronal fiber tracts also had a worse clinical presentation. This result demonstrates the importance of differential tractography as an important biomarker for disease progression in GM1 patients with potential extension to other neurodegenerative diseases and therapeutic intervention.

## Introduction

Diffusion weighed imaging (DWI) is a magnetic resonance imaging (MRI) technique utilizing multiple radio frequency pulses to evaluate water diffusion *in vivo*^1^. DWI approaches have expanded since the evolution of echo-planar imaging techniques, allowing for faster acquisition times^2,3^. DWI has proven to be useful in classifying cancer cells, ischemia, white matter diseases, and other conditions based on the extent of visualized local diffusion restriction^3–6^.

Diffusion tensor imaging (DTI) builds upon DWI by quantifying water’s local diffusion (isotropy) and diffusion restriction (anisotropy) utilizing the diffusion tensor matrix^7,8^. This quantification has given rise to classical DTI metrics, including fractional anisotropy (FA), mean diffusivity (MD), axial diffusivity (AD), and radial diffusivity (RD); these techniques assess axonal myelination and structural changes in the brain^9–11^. DTI has also allowed for the inception of fiber tractography, with the ability to map white matter neuronal pathways^12–14^.

Differential tractography is a new technique utilizing multiple longitudinal DTI scans with the capability of mapping alterations to specific white matter-derived neuronal pathways over time^15,16^. A classical DTI measure (FA/MD/AD/RD) is calculated for each fiber tract at baseline and follow-up timepoints, and the change is evaluated against a pre-determined threshold^15^. Fiber tracts exceeding that threshold are considered a growth or a loss depending on time orientation^15^.

Previous investigations utilizing differential tractography have been limited to adults with neuronal injury, with specific foci on Huntington’s disease^16^, multiple sclerosis^15^, end stage renal disease^17^, and traumatic brain injury^18^, demonstrating temporal changes associated with the degenerative progression of these conditions. To the best of our knowledge, no studies have been performed in children.

GM1 gangliosidosis, is an ultra-rare pan-ethnic degenerative neurological disease affecting 1 in 100,000-200,000 births^19^. GM1 gangliosidosis is caused by biallelic, loss-of-function mutations in *GLB1*, which encodes lysosomal β-galactosidase^20^. Decreased β-galactosidase activity results in the toxic accumulation of GM1 ganglioside and GA1 glycolipid, primarily in the central nervous system where the rate of synthesis of these molecules is highest^21,22^. GM1 clinically manifests as three types based on age at symptom onset and rate of disease progression^20^. GM1 Type I (infantile) has symptom onset in the first six months of life with rapid progression and death at 2-3 years^23^. GM1 Type II is divided into two subtypes, i.e., late-infantile with symptom onset between seven and twenty-four months and death in the second decade and juvenile, with onset at 3-5 years and survival into the 3^rd^ or 4^th^ decade^20,24^. GM1 Type III patients generally have symptom onset in the second decade and more gradual progression and clinical variability with long-term survival^22,25^.

There are no approved therapies for GM1 gangliosidosis and the disease is uniformly fatal^26^. However, adeno-associated virus (AAV) gene therapy techniques have been advancing rapidly and may be applicable to GM1 disease^27–30^. Objective outcome measures will be required to assess the efficacy of therapeutic interventions, such as AAV-mediated gene therapy, in GM1. DWI, whose use in GM1 has been limited to one case report that found hypo-intensities in the globus pallidum^31^, may fulfill this requirement. In this study, we present the first use of differential tractography to assess neuronal degeneration in children and adults with GM1 gangliosidosis compared with age-matched neurotypical developmental controls. The findings may be applicable as outcome imaging parameters to assess the efficacy of therapeutic interventions such as gene therapy.

## Methods

### The Natural History of GM1 Gangliosidosis

To determine the natural progression of GM1 gangliosidosis, participants from the NHGRI study, the “Natural History of Glycosphingolipid & Glycoprotein Storage Disorders” with a diagnosis of Type II GM1 diagnosis and repeated DWI were included in this analysis as the GM1 gangliosidosis cohort (NCT00029965)^32^. Ten GM1 natural history study (NHS) participants were included in the age-matched comparisons, and 16 GM1 NHS participants were included in the longitudinal analysis (see supplement A).

### Normal Controls

To determine how the GM1 cohort developed in relation to normal children, age-matched normal controls were included from two open-source data repositories. The OpenScienceFramework and OpenNeuro include the “Calgary Preschool magnetic resonance imaging (MRI) dataset”^33^ (n = 13) with participants aged 2-8 years and the “Queensland Twin Adolescent Brain (QTAB)”^34^ (n = 19) includes participants aged 9-16 years. Participants were selected for inclusion in this study by matching baseline age and latest follow up with the youngest GM1 patients (Figure 1).

**Figure 1.**
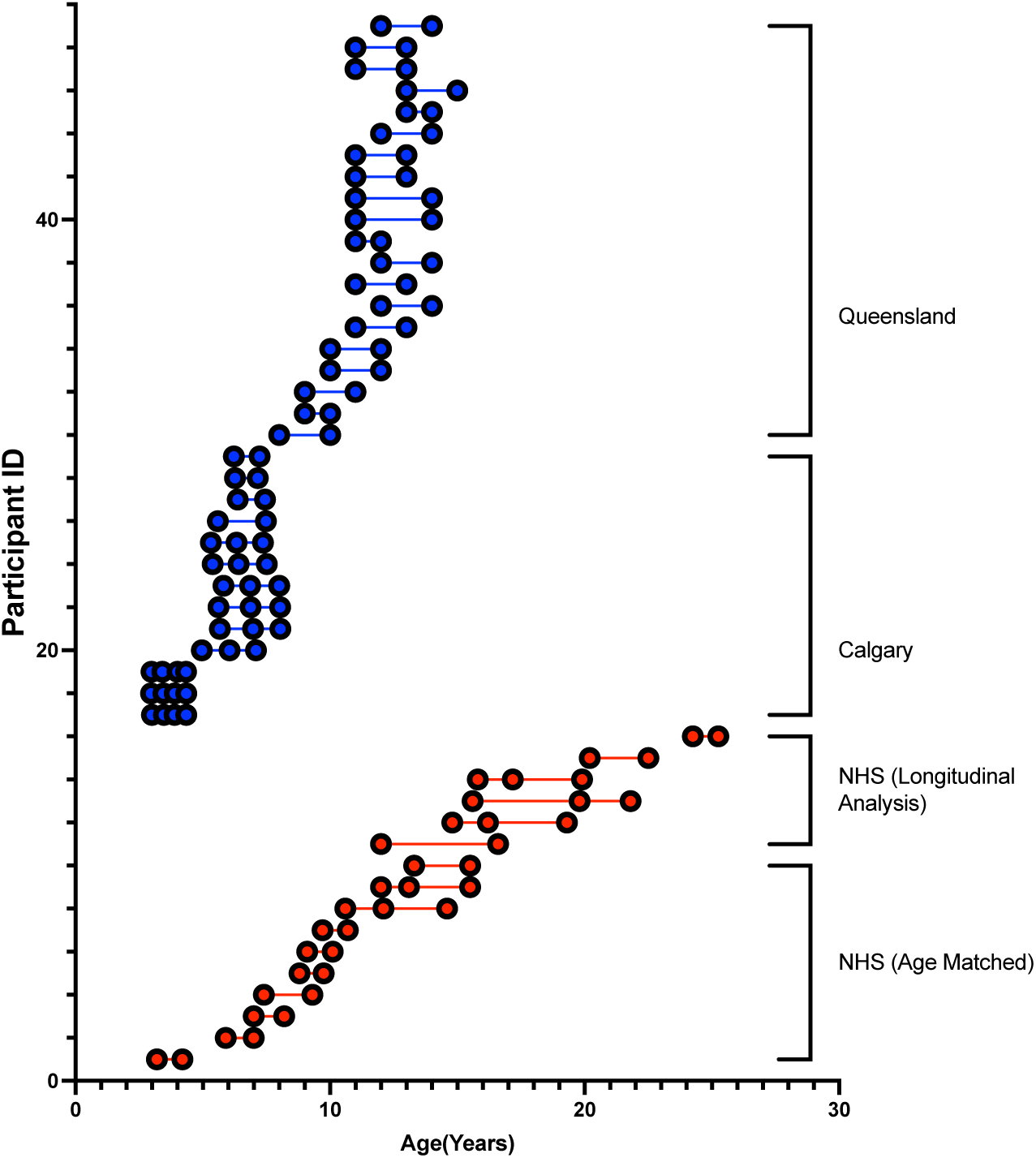
Participant age at each scan with GM1 patients shown in red and normal controls shown in blue. Each DWI scan is represented as a circle for all 113 scans where each of the 48 participants is on a separate row.

### CGI Scores

Cognitive Global Impression (CGI) is a clinician rating scale used to assess initial global illness severity (CGI-S) and overall change (CGI-C) from baseline with specific interventions^35^. It is extensively used in clinical trials and can be administered to a wide variety of patient populations. We retrospectively used CGI to assess our NHS patients at the beginning of the study (CGI-S) and longitudinally (CGI-C) during their participation in the study^36,37^.

### DWI Acquisition

#### Natural History Study Patients

NHS patients were sedated with propofol and/or sevoflurane for the duration of the scanning protocol. A Philips Achieva 3T system equipped with an 8-channel SENSE head coil was used to scan all NHS patients. DTI images were acquired with the following parameters: TR/TE=6400/100 ms, 32-gradient encoding directions, b-values=0 and 1000 s/mm^2^, voxel size=1.875mm×1.875mm×2.5mm, slice thickness=2.5 mm, acquisition matrix=128×128, NEX=1, FOV=24 cm.

### Calgary Normal Controls (NC)33

A General Electric 3T MR750w system with a 32-channel head coil was used for scanning all Calgary normal controls using a single shot spin echo-planar imaging sequence. DTI images were acquired with the following parameters for Calgary normal controls: TR/TE=6750/79 ms, 30-gradient encoding directions, b-values=0 and 750 s/mm^2^, voxel size=1.6mm×1.6mm×2.2mm, slice thickness=2.2 mm, FOV=20 cm.

### Queensland Normal Controls (NC)^34^

A 3T Magnetom Prisma (Siemens Medical Solutions, Erlangen) and a 64-channel head coil at the Centre for Advanced Imaging, University of Queensland employed a multi-shell with an anterior-posterior phase encoding direction. DTI images were acquired with the following parameters for Queensland normal controls: TR/TE= 3800/70 ms, 23-gradient encoding directions, b-values=0, 1,000, and 3,000 s/mm^2^, voxel size=2mm×2mm×2mm, slice thickness=2 mm, FOV=24 cm.

### DWI Processing

All DWI was preprocessed for artifacts, eddy currents, motion, and susceptibility induced distortions using MRtrix3’s (MRtrix, v3.0.4)^38^ *dwifslpreproc*^39–41^ command utilizing the *dwi2mask*^42^ function followed by FSL’s (FSL, v6.0.5) *eddy*^40^ and *topup*^40,41^ functions.

Preprocessed data were imported into DSI Studio (DSI Studio, v2023), where imaging was quality checked for bad slices, a U-Net mask was created, and generalized q-sampling imaging (GQI) based reconstruction was performed with a diffusion sampling length ratio of 1.25^43^ (see supplement B).

### Differential Tractography

Whole brain differential tractography was also performed in DSI Studio where fiber tract gains and losses were calculated using 10%, 20%, 30%, 40%, and 50% fractional anisotropy thresholds^15^. The angular threshold was 60 and the step size was 1 mm. Tracks < 20 mm or > 200 mm were discarded and 1,000,000 seeds were placed. Fiber tract gains were determined where the difference in FA between the follow-up and the baseline image exceeded the threshold. Fiber tract losses were determined where the difference in FA between the baseline and the follow-up image exceeded the threshold. Net fiber tract metrics were assessed as the difference between the growth and the loss.

To account for DWI sequencing and MRI scanner differences between cohorts, whole brain tractography was performed with the same parameters as above on each participant’s baseline diffusion weighted image. Percentage changes in fiber tract number and fiber tract volume were calculated relative to each participant’s baseline whole brain tractography.

### Statistical Analysis

Statistical analysis was performed in R studio (The R Foundation, v4.3.1). Between group analyses were performed between age-matched cohorts to demonstrate the effects of GM1 in 5 patients and 30 normal controls using Welch’s t-test. Longitudinal statistical analysis was performed between all participants who had serial DWI to demonstrate the viability of differential tractography as a biomarker and included 16 GM1 patients at 37 timepoints. Linear mixed effects modeling was used to evaluate longitudinal differential tractography parameters, the net percentage change in fiber tract number, and the net percentage change in fiber tract volume against CGI-C^44^.

## Results

### Age-matched Between Group Analysis

There were no significant differences in baseline age (*t* = 0.036, *p* = 0.9719), follow-up age (*t* = 0.036, *p* = 0.9839), or follow-up interval (*t* = 0.181, *p* = 0.8599) between the age-matched Natural History cohort and the normal controls (Figure 1).

Figure 2 shows substantial differences in longitudinal fiber tract development between GM1 patients and normal controls. At a low FA threshold (10%), the GM1 patients show drastic fiber tract losses (red) throughout the entire brain. In this patient, at a high FA threshold (50%) fiber tract losses are primarily located in the corpus callosum, a known location of abnormalities in GM1. In contrast, at a low FA threshold, the normal control shows significant fiber tract growth spread throughout the entire brain with minimal loss, while at a high FA (50%), there is milder and more localized growth with minimal fiber tract loss.

**Figure 2.**
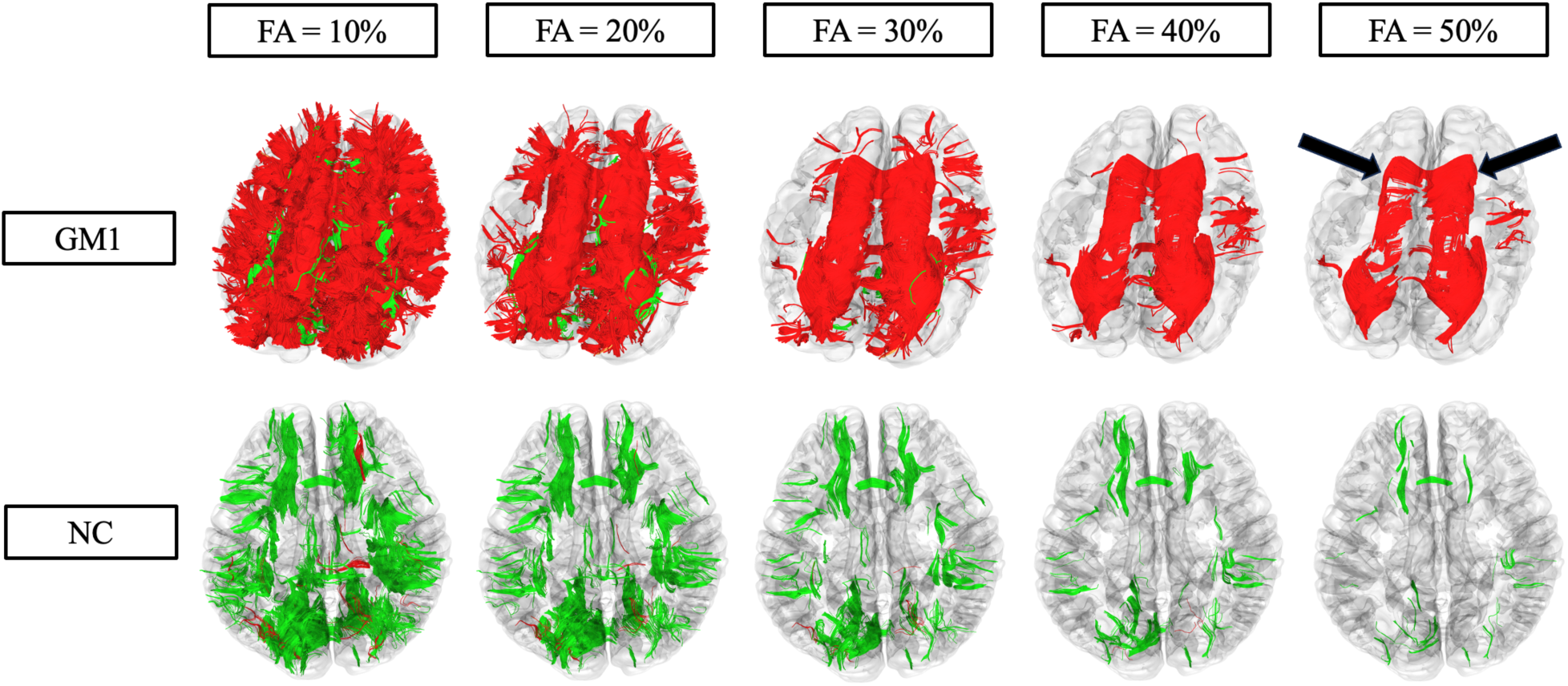
Differential tractography assessed fiber tract gains (green) and losses (red) at varying FA thresholds for one age matched late-infantile GM1 patient and one age matched normal control. At a low FA threshold (10%), the GM1 patient shows global and substantial fiber tract loss. At a high FA threshold (50%), the GM1 patients show milder fiber tract loss, localized primarily to the corpus callosum as indicated by the arrows. The neurotypical control shows global and moderate fiber tract growth at a low FA threshold (10%) with milder fiber tract growth at a high FA threshold (50%).

### Fiber Tractography Metrics

Normal controls showed statistically significant growth of fiber tract number and volume when compared to GM1 NHS patients at all FA thresholds (Figures 3&4). GM1 NHS patients showed statistically significant fiber tract number and volume loss when compared to normal controls at all FA thresholds (Figures 3&4).

**Figure 3.**
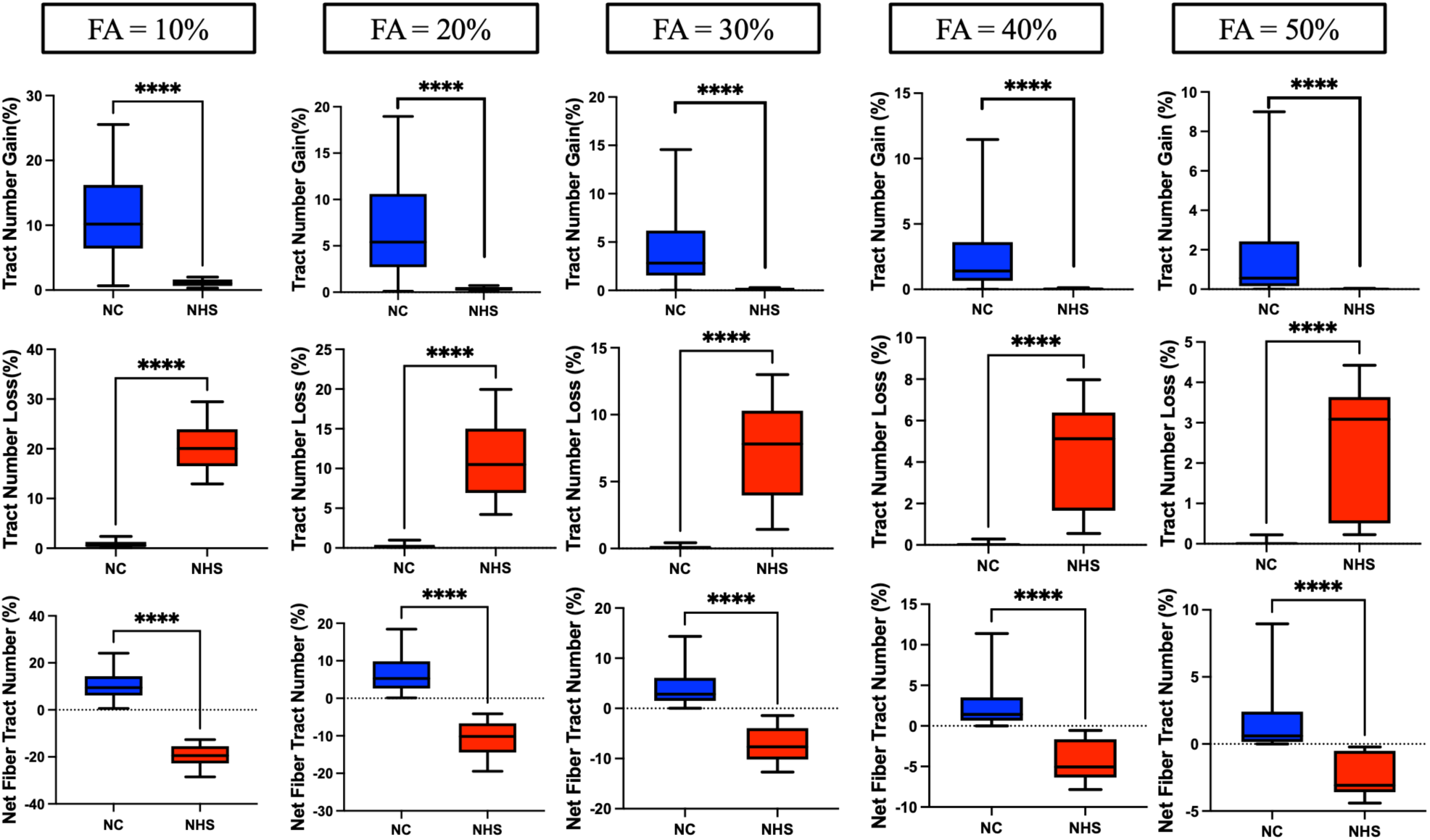
Age-matched differential tractography between group analysis of the number of fiber tracts. Row one indicates fiber tract growth as a percentage compared to baseline. Row two indicates fiber tract loss as a percentage compared to baseline. Row three indicates the net fiber tract number (growths minus losses) as a percentage compared to baseline. The columns indicate which fractional anisotropy threshold was tested from 10% to 50%.

**Figure 4.**
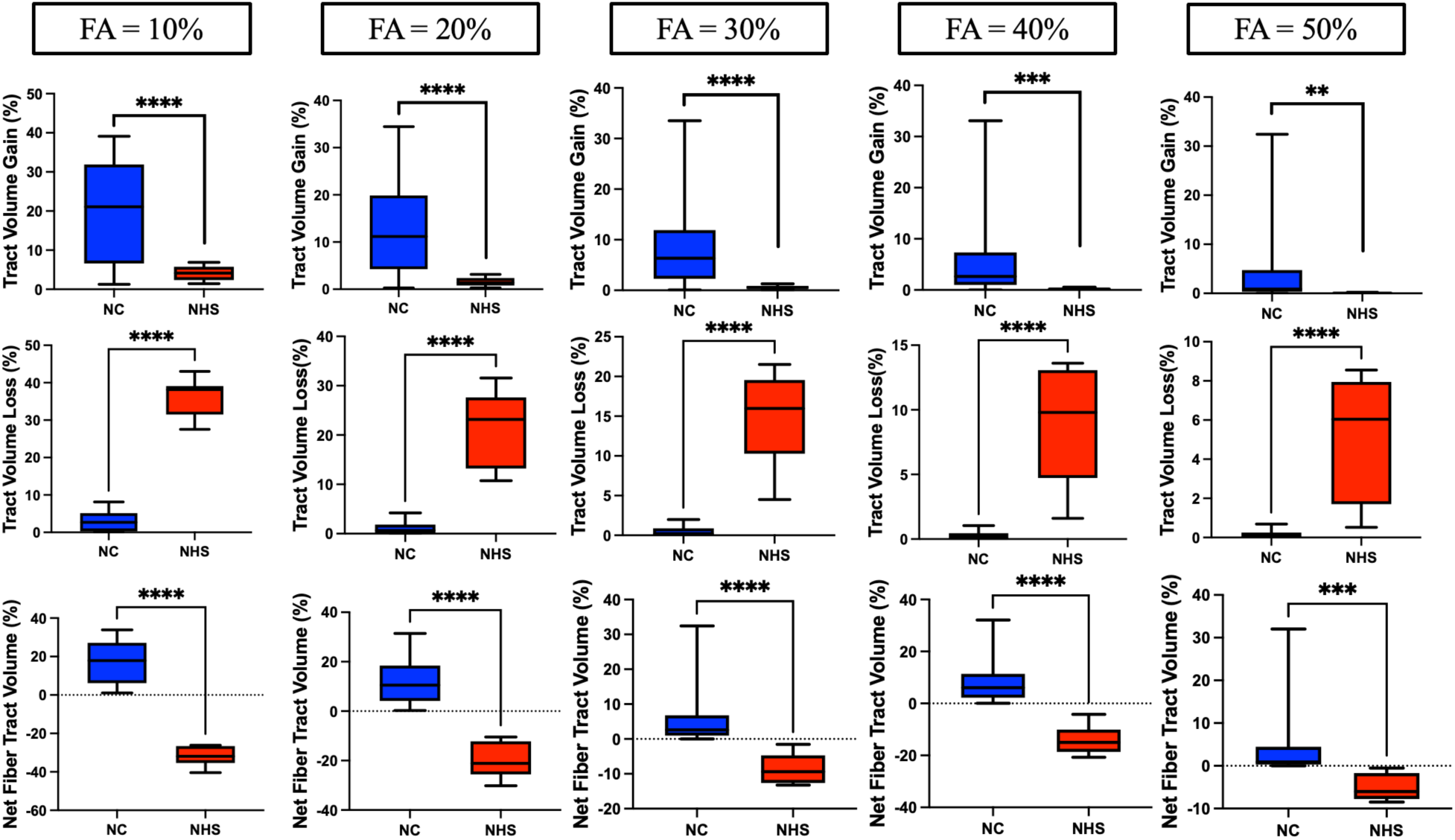
Age-matched differential tractography between group analysis of fiber tract volume. Row one indicates fiber tract volume increases as a percentage compared to baseline. Row two indicates fiber tract volume loss as a percentage compared to baseline. Row three indicates the net fiber tract volume (growth minus loss) as a percentage compared to baseline. The columns indicate which fractional anisotropy threshold was tested from 10% to 50%.

Figure 5 similarly demonstrates the longitudinal effects of GM1. GM1 patients show significant net fiber tract losses in both density (number) and size of fiber tracts (volume). Normal controls show growth in these domains associated with neurotypical development.

**Figure 5.**
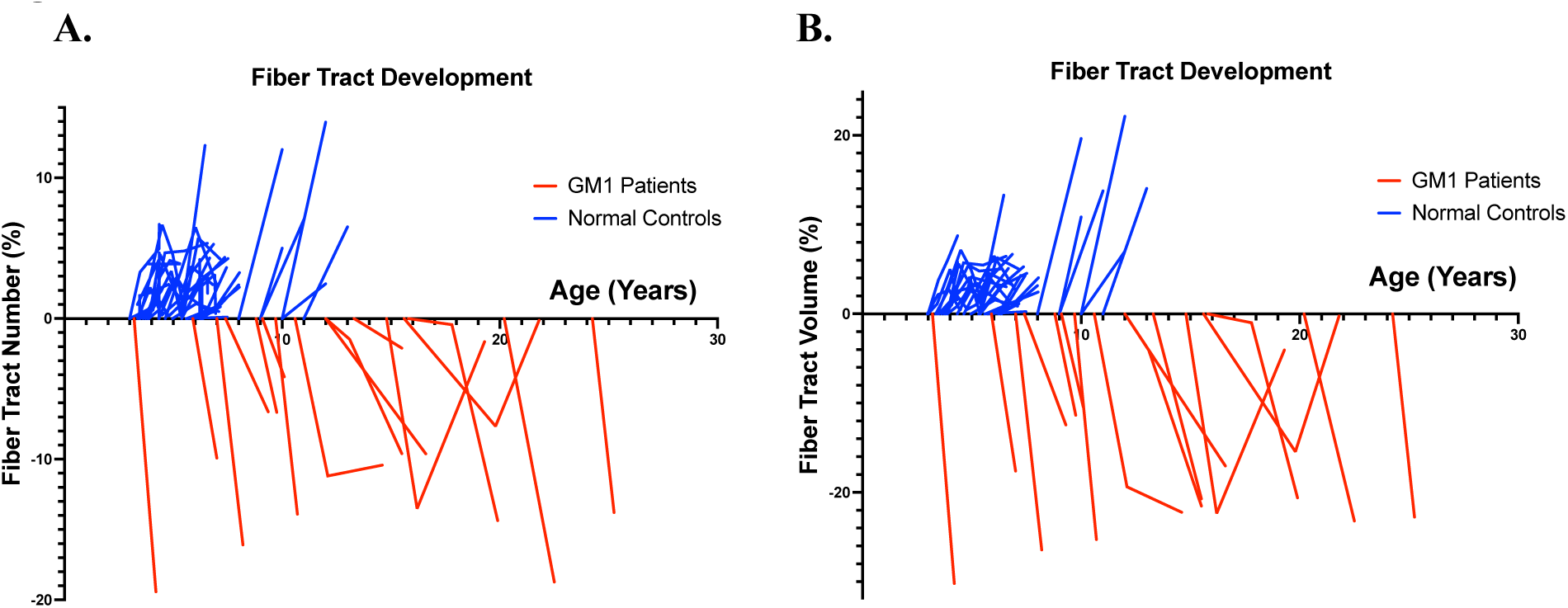
Differential tractography longitudinal analysis. A.) Net fiber tract number against participant age. B.) Net fiber tract volume against participant age.

### Longitudinal Analysis

Fiber tract number growth did not correlate with a change in clinical presentation as assessed by CGI-C (χ^2^ = 3.246, *p* = 0.0716, R^2^=0.0818). Fiber tract number loss (χ^2^ = 17.22, *p* < 0.001, R^2^=0.3655) and net fiber tract number (χ^2^ = 18.31, *p* < 0.001, R^2^ = 0.3837, Figure 6) both correlated with CGI-C. Fiber tract volume growth did not influence CGI-C (χ^2^ = 3.821, *p* = 0.0506, R^2^=0.0957). Fiber tract volume loss (χ^2^ = 23.01, *p* < 0.001, R^2^=0.4561) and net fiber tract volume (χ^2^ = 24.94, *p* < 0.001, R^2^ = 0.4833, Figure 6) both correlated with CGI-C.

**Figure 6.**
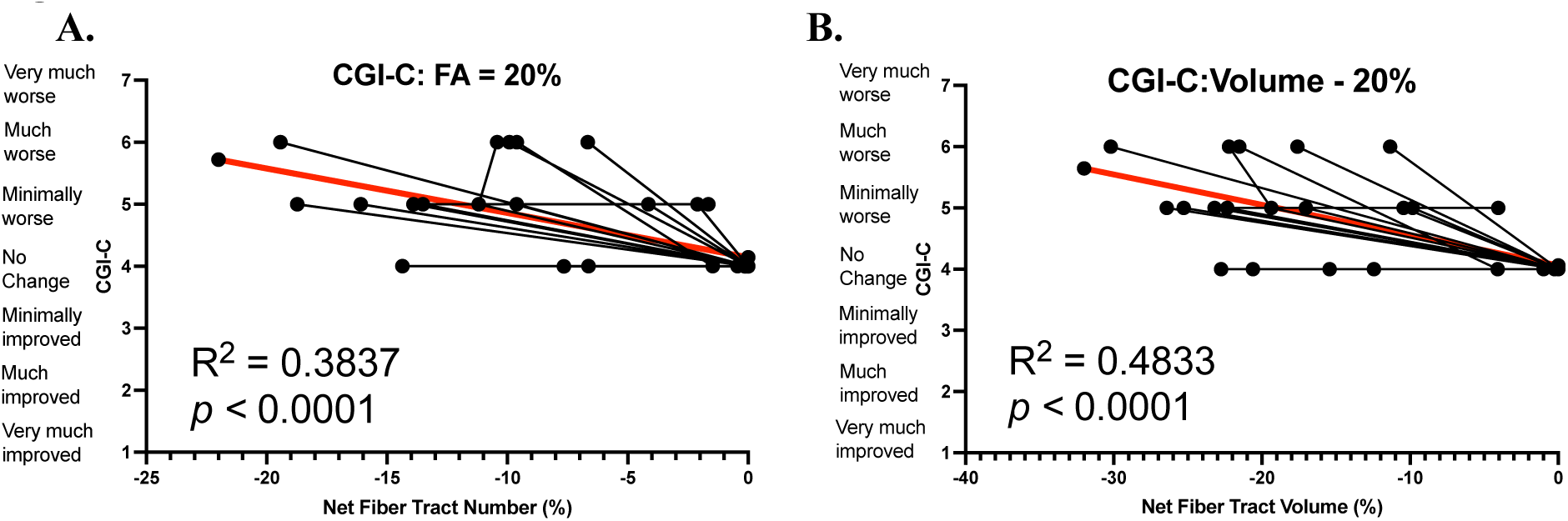
Differential Tractography correlations of net fiber tract number and net fiber tract volume with CGI-C change scores with GM1 patients at a 20% fractional anisotropy threshold.

## Discussion

In this study, we objectively identified and longitudinally quantified changes in fiber tract count and volume in normal individuals and in patients with GM1 gangliosidosis using differential tractography. This is a novel neuroradiological tool that allows the assessment of neuronal fiber track changes over time. We could infer that the changes reflect aberrations in neuronal cell growth resulting from neurodevelopmental and/or neurodegenerative conditions compared with neurotypical controls.

Previous investigations into differential tractography have been limited to adult degenerative disorders^15–18^. Here, we combined a population of children and adults with the same neurodegenerative condition (GM1) and compared their results with those of neurotypical individuals. This allowed for the assessment of not only the neurodegeneration associated with the medical condition, but also the expected gains and losses in typical neurodevelopmental trajectories.

Previous investigations into the white matter pathology of GM1 have demonstrated longitudinal degradation and delays in myelination or white matter development^45^. Our investigation supports these findings, since GM1 patients showed significant longitudinal neuronal cell loss with minimal growth in our age-matched analysis (Figure 2). Numerous studies^46–48^ have also demonstrated the neurotypical development of white matter through increases in FA and decreases in MD until reaching a peak at approximately 30 years of age. Our study also supports these findings, since net fiber tract metrics derived from FA changes were shown to increase in our neurotypical controls (Figure 5).

To better assess the importance and meaning of our neuroradiological findings related to brain structure and function, we correlated these results with clinical assessments used in our population to prepare for an upcoming clinical trial. The CGI-C is a widely used scale in clinical trials that allows the assessment of clinical changes associated with tested interventions. It is traditionally used as a prospective assessment in addition to other outcome measurements selected for a specific study. In our case, we used a novel retrospective version of the CGI to assess disease progression over time from historical records^37^. In our longitudinal analysis of differential tractography, we found significant correlations between net differential tractography metrics and clinical presentation assessed by CGI-C; increased fiber tract loss corresponded to a worsening in the clinical presentation (Figure 6). This demonstrates the utility of differential tractography as a biomarker in evaluating longitudinal change in the clinical presentation of patients with GM1 and potentially other neurodegenerative diseases.

To determine the sensitivity of these results, we tested our fiber tract metrics at varying FA thresholds between 10 and 50 percent. Only fiber tracts with a change in FA exceeding the specified threshold in both directions (growth and loss) were identified by differential tractography. We found significant differences in growth and loss of both the number of fiber tracts and the volume of these tracts at all FA thresholds between GM1 gangliosidosis patients and neurotypical controls. This suggests that our results are independent of the FA threshold being examined and demonstrates the utility of differential tractography in differentiating between neurotypical and neurodegenerative developmental white matter changes. While a 10% FA threshold exhibited stronger results in terms of fiber tract growth and loss (Figure 3&4), it likely also had more false discoveries as described in Yeh et al^15^. Our longitudinal analysis also supported this notion; at all FA thresholds between 10% and 50%, we found statistically significant correlations between net fiber tract metrics and CGI-C scores (see supplement C). Lower FA thresholds yielded stronger correlations with CGI-C; this requires further validation.

Some of the limitations in our study need to be considered as we aim to use this methodology in clinical trials and other research projects that require identification of temporal changes in brain anatomy and structure. One limitation is the variability in scanning sequences among GM1 patients and normal controls. We think this issue was mitigated by using the baseline whole brain tractography, which allows for the relative percentage of fiber tract growth and loss for each participant to be calculated prior to inter-cohort comparisons. Similarly, the smaller number of diffusion directions in the scanning sequences is a limitation of this study. However, while Yeh et al^15^. demonstrated the limitations associated with a reduced number of diffusion directions, they found this limitation is associated with a smaller number of detections; this suggests that the use of an optimal scanning protocol could yield even more impressive differential tractography results. Another limitation of this study is the absence of a sham sequence and subsequent analysis of the false discovery rate^15^. GM1 NHS participants also underwent propofol sedation during DWI acquisition where neurotypical controls remained awake during their DWI acquisition protocols. Previous studies have suggested that propofol does not influence DTI parameters^49,50^, but this requires further validation. Lastly, this study is limited by a small sample size; future studies evaluating the role of differential tractography as a neurogenerative biomarker should incorporate a larger sample size to demonstrate reliability. Nevertheless, we believe that this work represents an important step forward in the identification of biomarkers for disease progression that considers the well-known but rarely included changes in brain structure and function associated with neurodevelopment and aging.

## Conclusion

This study is the first to explore the utility of differential tractography in demonstrating longitudinal changes in white matter fiber tracts resulting from neurodevelopment and neurodegeneration in GM1 gangliosidosis. Overall, GM1 patients showed statistically significant loss of white matter tract count and white matter tract volume, reflecting the natural progression of GM1. Differential tractography results strongly correlated with longitudinal clinical outcomes as measured by CGI-C in GM1 gangliosidosis patients. These results indicate the importance of differential tractography as a robust biomarker for disease progression in GM1 patients, and potentially extend to a similar role in other neurodegenerative diseases and lysosomal storage disorders.

## Data Availability

The data described in this manuscript are available from the corresponding author upon reasonable request.

## Acknowledgments

We thank the participants and their families for the generosity of their time and efforts. We are also grateful to many staff members and care providers who contributed their expertise over the years. Magnetic resonance imaging analysis in this work utilized the computational resources of the Biowulf Linux cluster at the National Institutes of Health (http://hpc.nih.gov).

## Author Contributions

Conceptualization: CJL, MSS, MTA, CJT; Data Curation: PD, JMJ, CJT, MTA; Funding Acquisition: CJT; Methodology: CJL, ZV, MSS, CJT, MTA; Visualization: CJL, MTA; Writing-original draft: CJL, WAG, CJT, MTA; Writing-reviewing & editing; ZV, ALK, PD, JMJ, WAG, MSS, CJT, MTA

## Ethics Declaration

The NIH Institutional Review Board approved this protocol (02-HG-0107). Informed consent was completed with parents or legal guardians of the patients. All participants were assessed for their ability to provide assent; none were deemed capable.

## Supplementary Methods

### Supplement A: Natural History Study Age-Matched Methodology

#### NHGRI Natural History Study (NCT00029965)^1^

##### Study description

This is a natural history study that will evaluate any patient with enzyme-or DNA-confirmed GM1 or GM2 gangliosidosis, sialidosis or galactosialidosis. Patients may be evaluated every 6 months for infantile onset disease, yearly for juvenile onset and approximately every two years for adult-onset disease as long as they are clinically stable to travel. Data will be evaluated serially for each patient and cross-sectionally for patients of similar ages and genotypes.

Genotype-phenotype correlations will be made where possible although these are rare disorders and the majority of the patients are compound heterozygotes.

##### Objectives

– To study the natural history and progression of neurodegeneration in individuals with glycosphingolipid storage disorders (GSL), GM1 and GM2 gangliosidosis, and glycoprotein (GP) disorders including sialidosis and galactosialidosis using clinical evaluation of patients and patient/parent surveys.
– To develop sensitive tools for monitoring disease progression.
– To identify biological markers in blood, cerebrospinal fluid, and urine that correlate with disease severity and progression and can be used as outcome measures for future clinical trials.
– To further understand and characterize the mechanisms of neurodegeneration in GSL and GP storage disorders across the spectrum of disease beginning with ganglioside storage in fetal life.

##### Study Population

Patients with enzyme-or DNA-confirmed GM1 or GM2 gangliosidosis, sialidosis or galactosialidosis. Accrual ceiling is 200 participants, with no exclusions based on age, gender, demographic group, or demographic location. Patients included in our study are those who were seen at the NIH Clinical Center or who only sent in blood samples or who complete the questionnaire or provided head circumference measures.

##### Inclusion Criteria

– Individuals greater than 6 months of age with GM1 or GM2 gangliosidosis documented by enzyme deficiency and/or mutation analysis in a CLIA-approved laboratory

##### Exclusion Criteria

– Individuals who in the opinion of the principal investigator are too medically fragile to travel safely to the NIH for evaluation
– Individuals unable to comply with the protocol

### NHGRI Natural History Study Age Matched Cohort

The data included in this investigation represents a subset of the natural history study patients. This study includes only patients who had a confirmed GM1 Gangliosidosis diagnosis, excluding those with other glycosphingolipid storage disorders, glycoprotein disorders, and GM2 Gangliosidosis who were a part of the larger natural history cohort (**Table A1**). Patients were selected for the longitudinal analysis cohort based on having multiple diffusion weighted imaging scans with corresponding cognitive global impression (CGI) scores (**Table A2**). Patients were selected for the age-matched cohort based on their baseline scan age and follow-up scan (table). Only patients who had repeated diffusion weighted imaging scans within the range of the normal controls (2.5 years old – 16 years old) were included (**Table A3**).

**Table A1.**
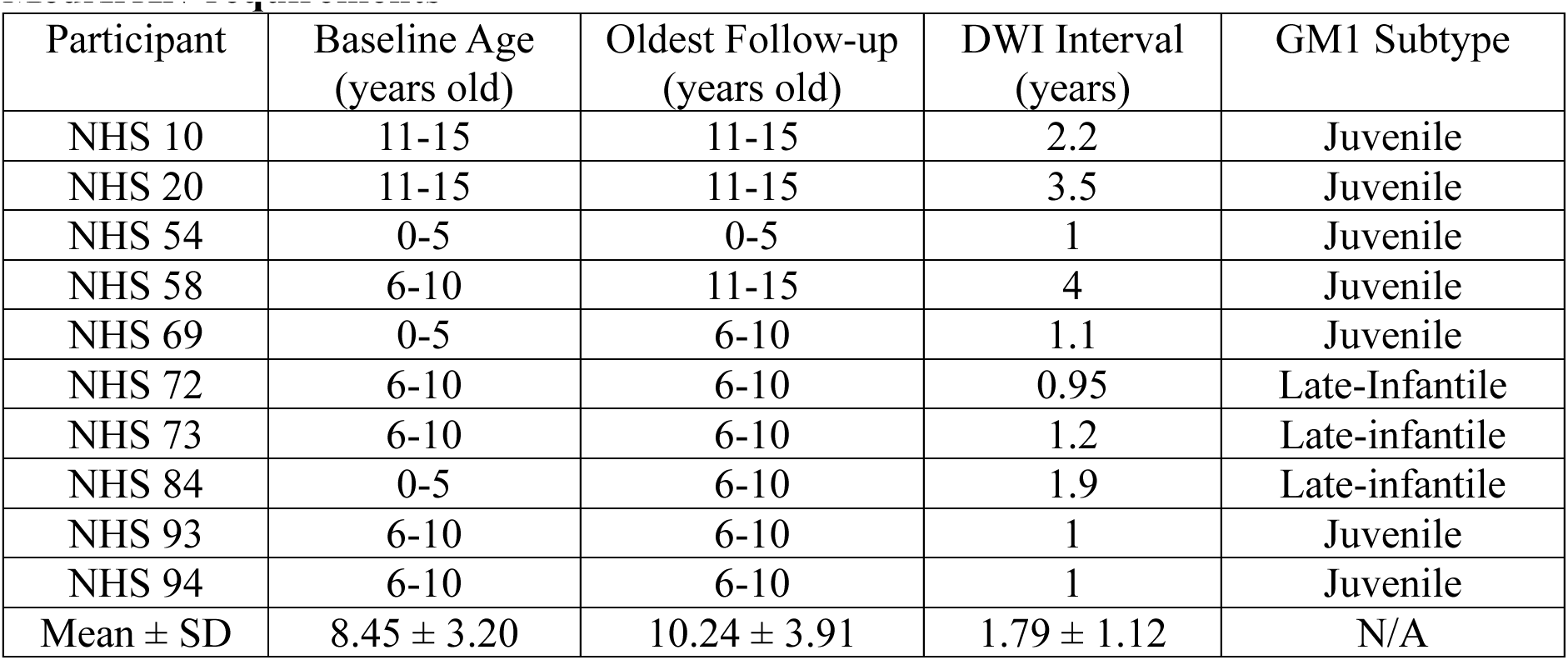
Natural History Study Age Matched Cohort (n = 10), specific ages redacted per MedArXiv requirements.

| Participant | Baseline Age (years old) | Oldest Follow-up (years old) | DWI Interval (years) | GM1 Subtype |
| --- | --- | --- | --- | --- |
| NHS 10 | 11-15 | 11-15 | 2.2 | Juvenile |
| NHS 20 | 11-15 | 11-15 | 3.5 | Juvenile |
| NHS 54 | 0-5 | 0-5 | 1 | Juvenile |
| NHS 58 | 6-10 | 11-15 | 4 | Juvenile |
| NHS 69 | 0-5 | 6-10 | 1.1 | Juvenile |
| NHS 72 | 6-10 | 6-10 | 0.95 | Late-Infantile |
| NHS 73 | 6-10 | 6-10 | 1.2 | Late-infantile |
| NHS 84 | 0-5 | 6-10 | 1.9 | Late-infantile |
| NHS 93 | 6-10 | 6-10 | 1 | Juvenile |
| NHS 94 | 6-10 | 6-10 | 1 | Juvenile |
| Mean ± SD | 8.45 ± 3.20 | 10.24 ± 3.91 | 1.79 ± 1.12 | N/A |

**Table A2.** Natural History Study Longitudinal Analysis Cohort (n = 16), specific ages redacted per MedArXiv requirements.

| Participant | GM1 Sub-type | Baseline Age (years old) | Scan #2 Age (years old) | Scan #3 Age (years old) | Average DWI Interval (years) | Number of DWI Scans |
| --- | --- | --- | --- | --- | --- | --- |
| NHS 03 | Juv | 21-25 | 21-25 | N/A | 1.0 | 2 |
| NHS 09 | Juv | 11-15 | 16-20 | 21-25 | 3.1 | 3 |
| NHS 10 | Juv | 11-15 | 11-15 | N/A | 2.2 | 2 |
| NHS 11 | Juv | 11-15 | 16-20 | 16-20 | 2.05 | 3 |
| NHS 20 | Juv | 11-15 | 11-15 | 11-15 | 1.75 | 3 |
| NHS 25 | Juv | 11-15 | 16-20 | 16-20 | 2.25 | 3 |
| NHS 26 | Juv | 11-15 | 16-20 | N/A | 4.6 | 2 |
| NHS 28 | Juv | 16-20 | 21-25 | N/A | 2.3 | 2 |
| NHS 54 | Juv | 0-5 | 0-5 | N/A | 1.0 | 2 |
| NHS 58 | Juv | 5-10 | 11-15 | 11-15 | 2.0 | 3 |
| NHS 69 | Juv | 0-5 | 6-10 | N/A | 1.1 | 2 |
| NHS 72 | LI | 6-10 | 6-10 | N/A | 0.95 | 2 |
| NHS 73 | LI | 6-10 | 6-10 | N/A | 1.2 | 2 |
| NHS 84 | LI | 0-5 | 6-10 | N/A | 1.9 | 2 |
| NHS 93 | Juv | 6-10 | 6-10 | N/A | 1.0 | 2 |
| NHS 94 | Juv | 6-10 | 6-10 | N/A | 1.0 | 2 |

**Table A3.** Normal Control Age Matched Cohort (n = 32), specific ages redacted per MedArXiv requirements.

| Participant | Baseline Age<br>(years old) | Oldest Follow-up<br>(years old) | DWI Interval<br>(years) | Database |
| --- | --- | --- | --- | --- |
| 10073 | 0-5 | 0-5 | 1.3528 | Calgary |
| 10007 | 0-5 | 0-5 | 1.3528 | Calgary |
| 10066 | 0-5 | 0-5 | 1.39 | Calgary |
| 10054 | 0-5 | 6-10 | 2.4163 | Calgary |
| 10148 | 0-5 | 6-10 | 2.1889 | Calgary |
| 10109 | 0-5 | 6-10 | 2.1306 | Calgary |
| 10022 | 0-5 | 6-10 | 2.37334 | Calgary |
| 10025 | 0-5 | 6-10 | 2.05 | Calgary |
| 10027 | 0-5 | 6-10 | 1.8889 | Calgary |
| 10090 | 0-5 | 6-10 | 2.1278 | Calgary |
| 10020 | 6-10 | 6-10 | 1.075 | Calgary |
| 10087 | 6-10 | 6-10 | 0.8889 | Calgary |
| 10161 | 6-10 | 6-10 | 1.0166 | Calgary |
| 360 | 6-10 | 11-15 | 2 | QTAB |
| 410 | 11-15 | 11-15 | 2 | QTAB |
| 411 | 6-10 | 11-15 | 2 | QTAB |
| 376 | 6-10 | 6-10 | 1 | QTAB |
| 378 | 6-10 | 11-15 | 2 | QTAB |
| 405 | 6-10 | 6-10 | 2 | QTAB |
| 200 | 11-15 | 11-15 | 2 | QTAB |
| 197 | 11-15 | 11-15 | 2 | QTAB |
| 190 | 11-15 | 11-15 | 1 | QTAB |
| 186 | 11-15 | 11-15 | 3 | QTAB |
| 183 | 11-15 | 11-15 | 3 | QTAB |
| 172 | 11-15 | 11-15 | 2 | QTAB |
| 173 | 11-15 | 11-15 | 2 | QTAB |
| 162 | 11-15 | 11-15 | 2 | QTAB |
| 158 | 11-15 | 11-15 | 1 | QTAB |
| 152 | 11-15 | 11-15 | 2 | QTAB |
| 157 | 11-15 | 11-15 | 2 | QTAB |
| 156 | 11-15 | 11-15 | 2 | QTAB |
| 155 | 11-15 | 11-15 | 2 | QTAB |
| Mean ± SD | 8.54 ± 3.16 | 10.39 ± 3.32 | 1.85 ± 0.54 | N/A |

### Supplement B: Diffusion Weighted Imaging (DWI) Sequence Parameters and Processing

#### Natural History Study (NHS) Patients^1^

A Philips Achieva 3T system equipped with an 8-channel SENSE head coil was used to scan all Natural History Study patients. DTI images were acquired with the following parameters for NHS: TR/TE=6400/100 ms, 32-gradient directions, b-values=0 and 1000 s/mm^2^, slice thickness=2.5 mm, acquisition matrix=128×128, NEX=1, FOV=24 cm.

#### Calgary Normal Controls (NC)^2^

A General Electric 3T MR750w system and a 32-channel head coil was used for scanning all Calgary normal controls using a single shot spin echo-planar imaging sequence. DTI images were acquired with the following parameters for Calgary normal controls: TR/TE=6750/79 ms, FOV=20 cm, 30 gradient encoding directions at b=0 and 750 s/mm^2^.

#### Queensland Normal Controls(NC)^3^

A 3T Magnetom Prisma (Siemens Medical Solutions, Erlangen) and a 64-channel head coil at the Centre for Advanced Imaging, University of Queensland using a multi-shell with an anterior-posterior phase encoding direction. DTI images were acquired with the following parameters for Queensland normal controls: TR/TE= 3800/70 ms, voxel size=2mm x 2mm x 2mm,23-gradient directions, b-values=0, 1,000, and 3,000 s/mm^2^, slice thickness=2 mm, FOV=244×244mm.

#### DWI Preprocessing (*Fig. B1*)

DWI was first converted from DICOM to a NIFTI file using *dcm2niix* where the b-values and b-vectors files were acquired^4^. DWI at all timestamps was preprocessed for artifacts, eddy currents, motion, and susceptibility induced distortions using MRtrix3’s (MRtrix, v3.0.4)^5^ *dwifslpreproc*^6–8^ command utilizing the *dwi2mask*^9^ function followed by FSL’s (FSL, v6.0.5) *eddy*^7^ and *topup*^7,8^ functions. Preprocessed data was imported into DSI Studio (DSI Studio, v2023) where imaging was quality checked for bad slices, a U-Net mask was created, and generalized q-sampling imaging (GQI) reconstruction was performed with a diffusion sampling length ratio of 1.25^10^.

#### DWI Processing (*Fig. B2*)

First, the fractional anisotropy (FA) map of the baseline image was exported as a NIFTI file. Whole brain fiber tractography was then performed on the baseline image with 1,000,000 seeds, a step size of 1 mm, an angular threshold of 60, minimum tract size of 20 mm, and a maximum tract size of 200 mm. Differential tractography was then performed on each subsequent follow-up scan in comparison with the baseline image where fiber tract gains and losses were calculated using 10%, 20%, 30%, 40%, and 50% fractional anisotropy thresholds. Fiber tract gains were determined where the difference in FA between the follow-up and the baseline image exceeded the threshold 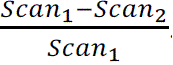. Fiber tract losses were determined where the difference in FA between the baseline and the follow-up image exceeded the threshold utilizing the equation 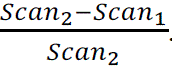. Differential tractography was calculated with the following parameters: angular threshold=60, step size=1 mm, tracts < 20 mm or > 200 mm were discarded, and 1,000,000 seeds were placed.

**Figure B1.**
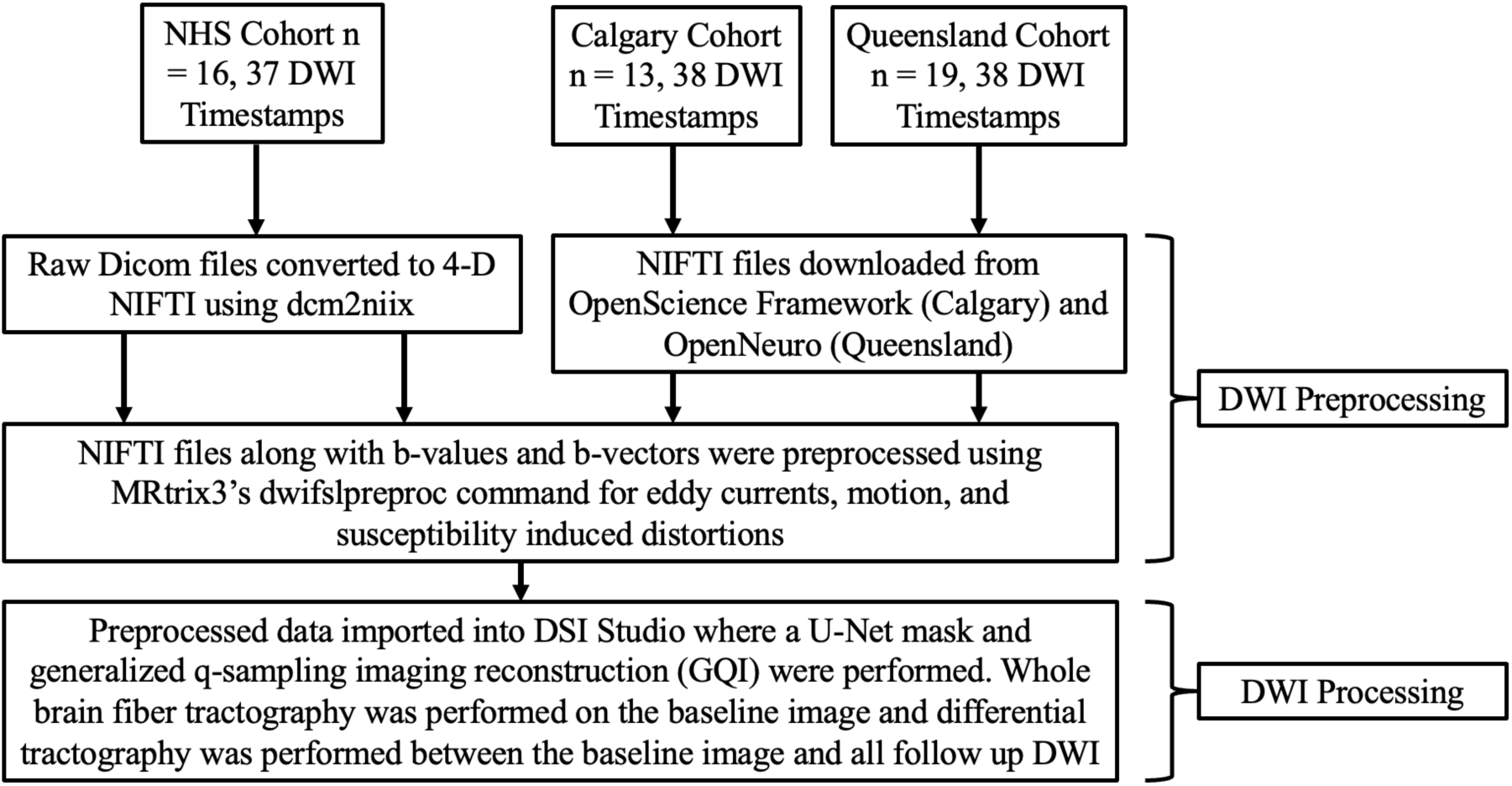
DWI preprocessing pipeline.

**Figure B2.**
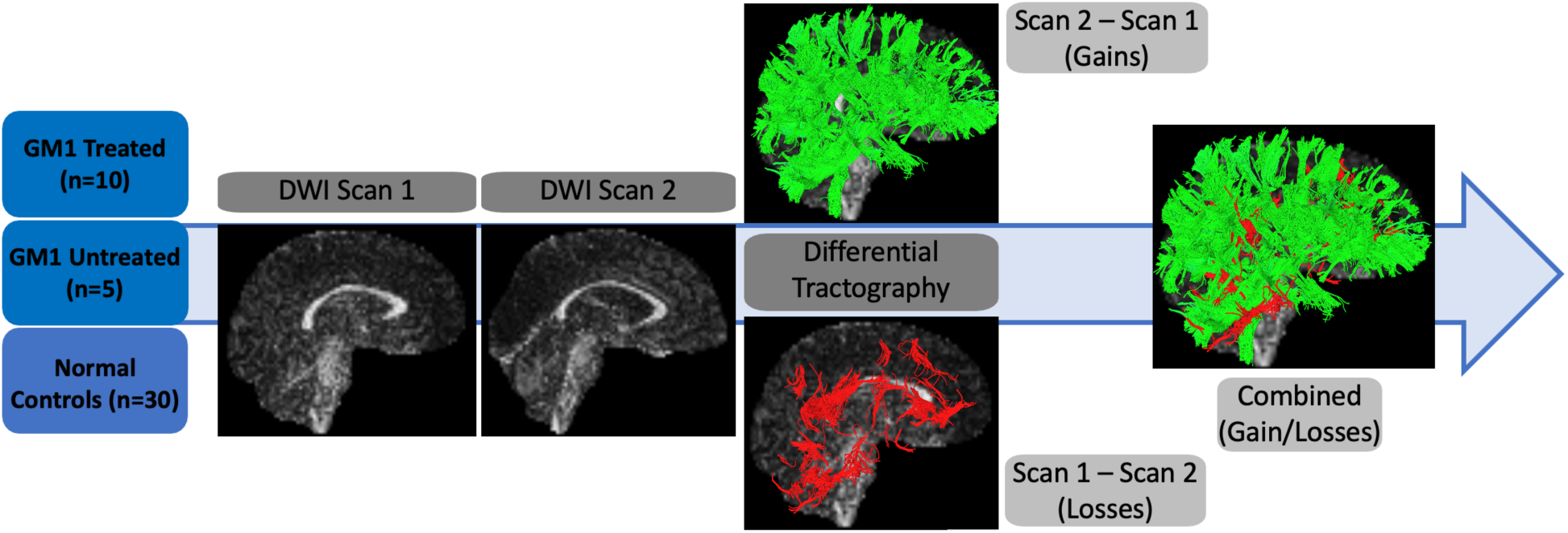
Differential Tractography overview.

## Supplementary Results

### Supplement C: FA Thresholds on CGI-C

**Figure C1.**
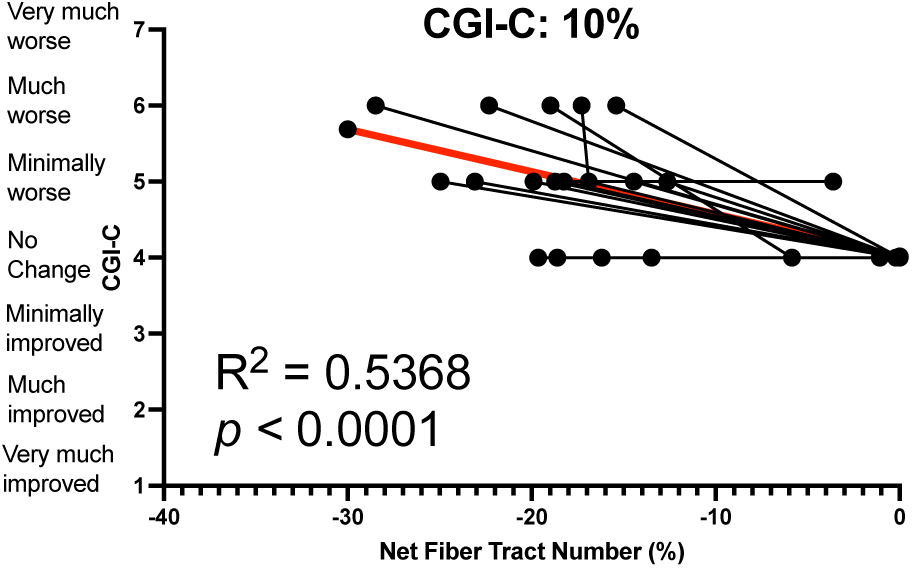
Differential Tractography correlations of net fiber tract number with CGI-C change scores with GM1 patients at a 10% fractional anisotropy threshold.

**Figure C2.**
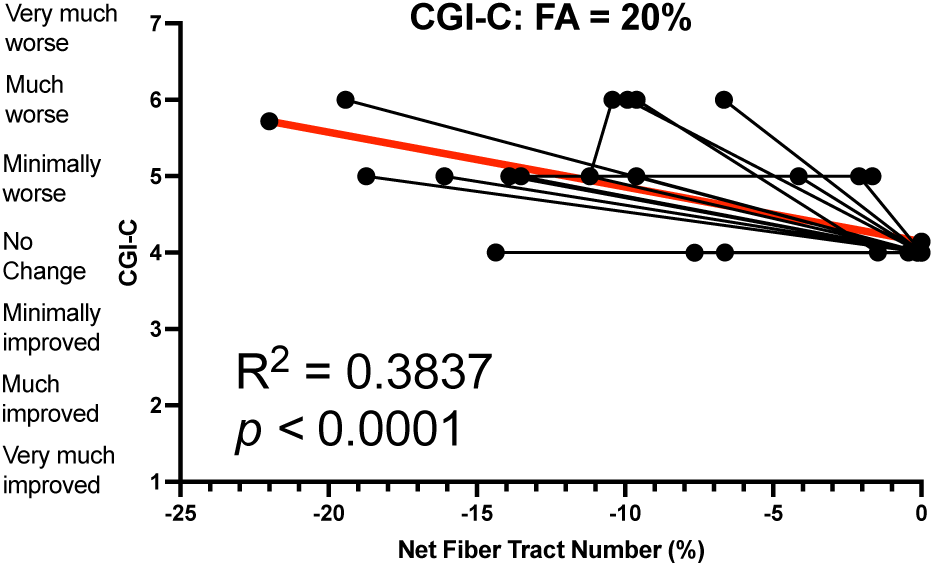
Differential Tractography correlations of net fiber tract number with CGI-C change scores with GM1 patients at a 20% fractional anisotropy threshold.

**Figure C3.**
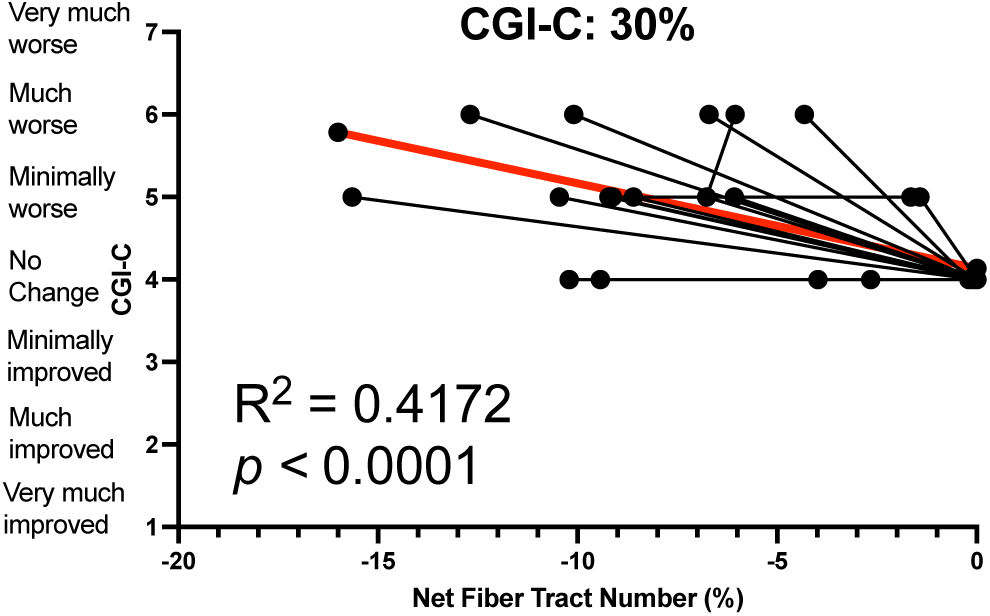
Differential Tractography correlations of net fiber tract number with CGI-C change scores with GM1 patients at a 30% fractional anisotropy threshold.

**Figure C4.**
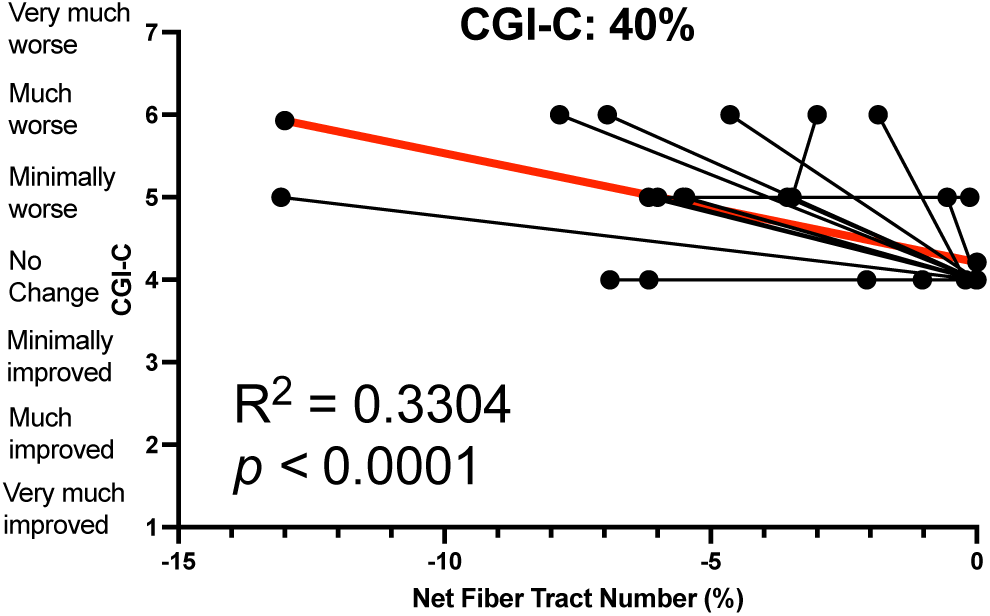
Differential Tractography correlations of net fiber tract number with CGI-C change scores with GM1 patients at a 40% fractional anisotropy threshold.

**Figure C5.**
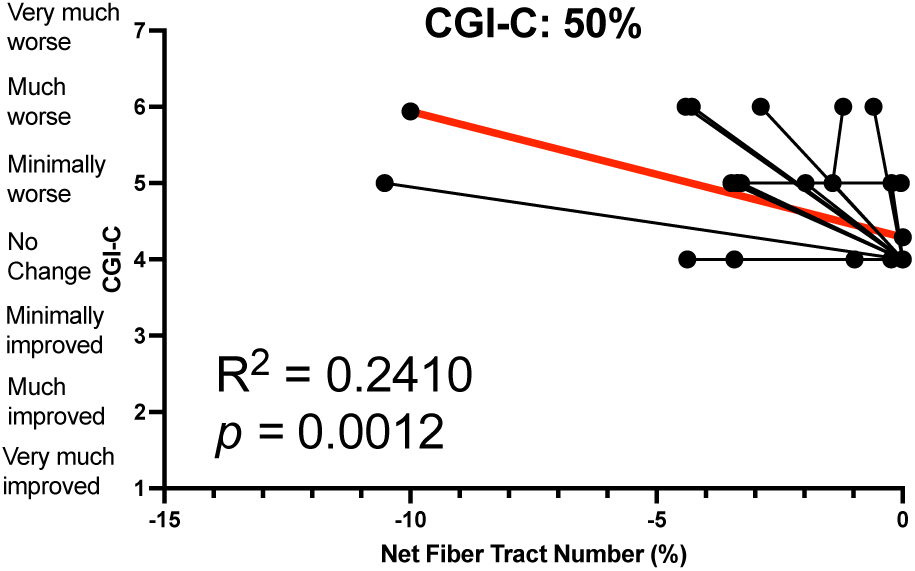
Differential Tractography correlations of net fiber tract number with CGI-C change scores with GM1 patients at a 50% fractional anisotropy threshold.

**Figure C6.**
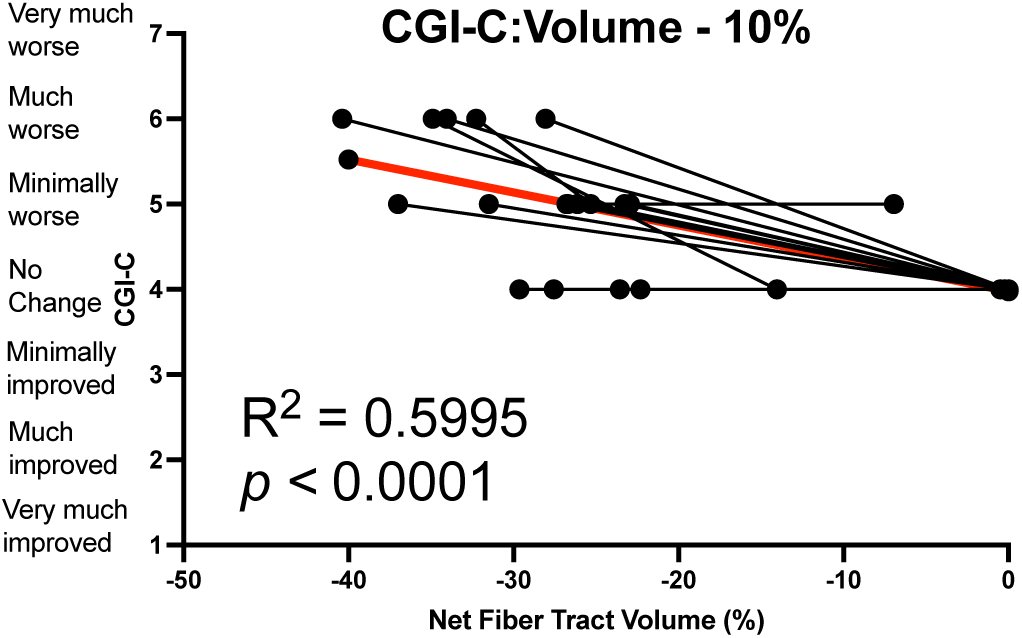
Differential Tractography correlations of net fiber tract volume with CGI-C change scores with GM1 patients at a 10% fractional anisotropy threshold.

**Figure C7.**
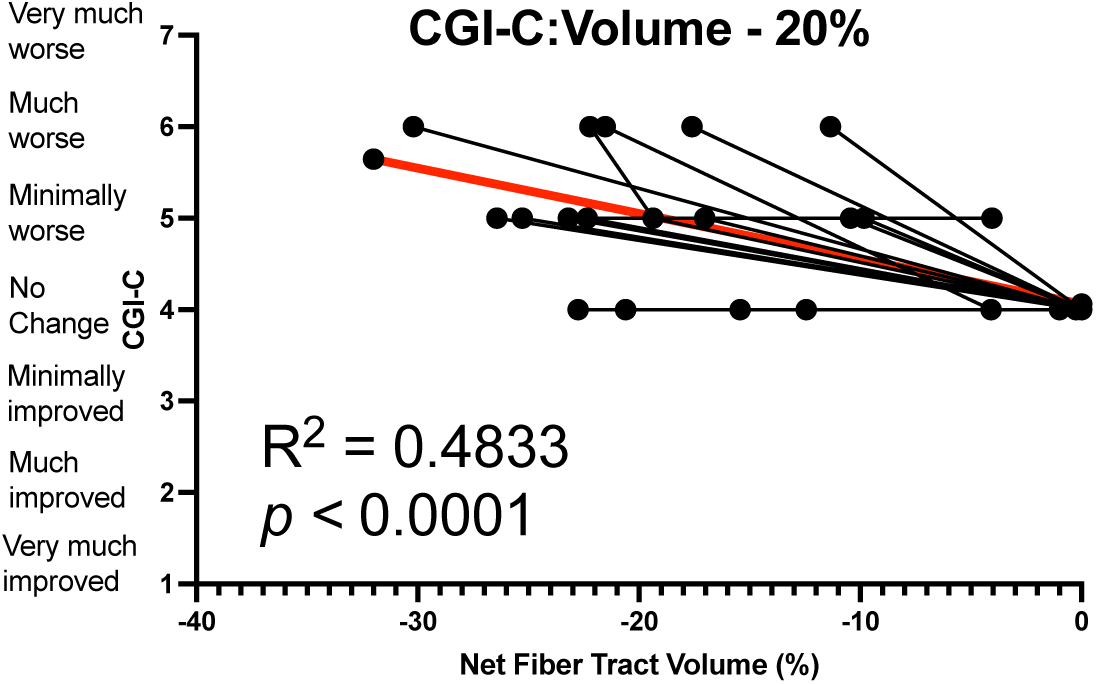
Differential Tractography correlations of net fiber tract volume with CGI-C change scores with GM1 patients at a 20% fractional anisotropy threshold.

**Figure C8.**
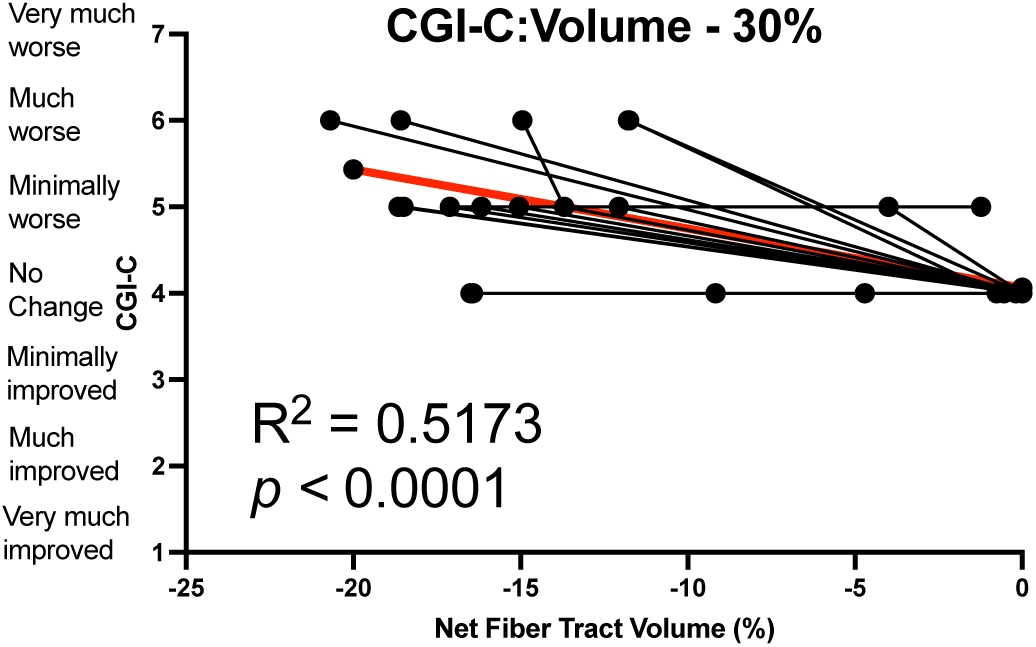
Differential Tractography correlations of net fiber tract volume with CGI-C change scores with GM1 patients at a 30% fractional anisotropy threshold.

**Figure C9.**
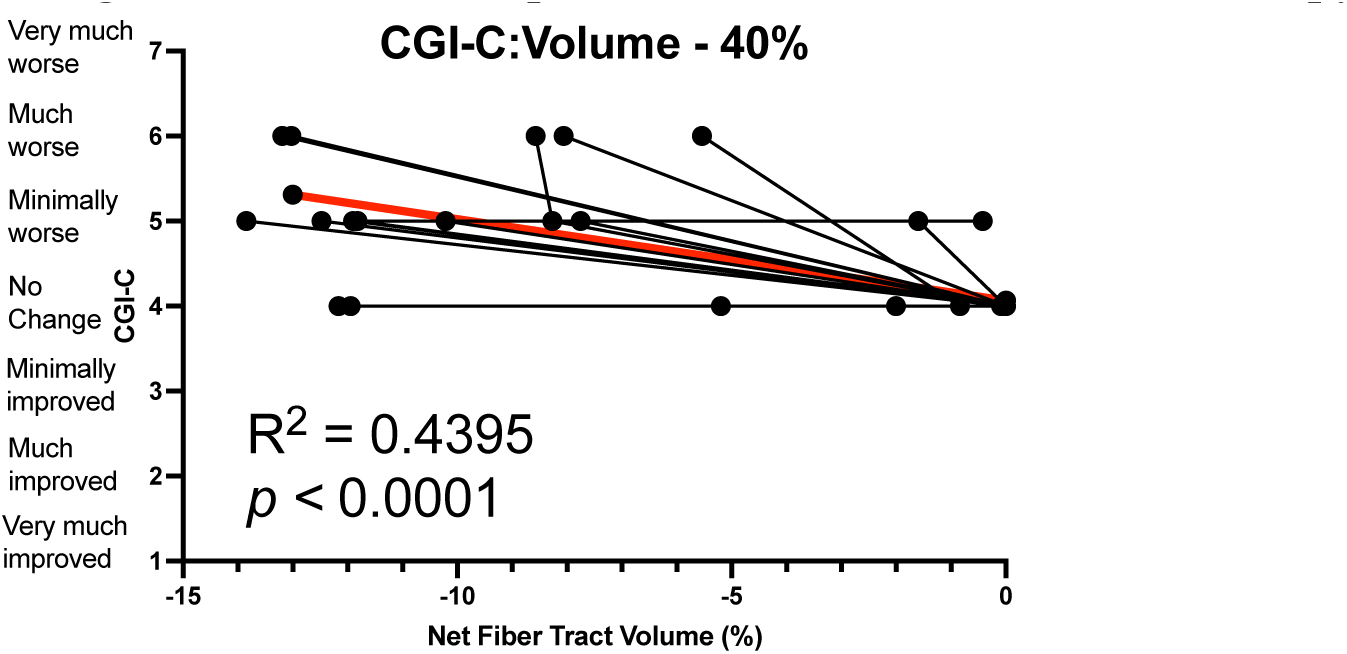
Differential Tractography correlations of net fiber tract volume with CGI-C change scores with GM1 patients at a 40% fractional anisotropy threshold.

**Figure C10.**
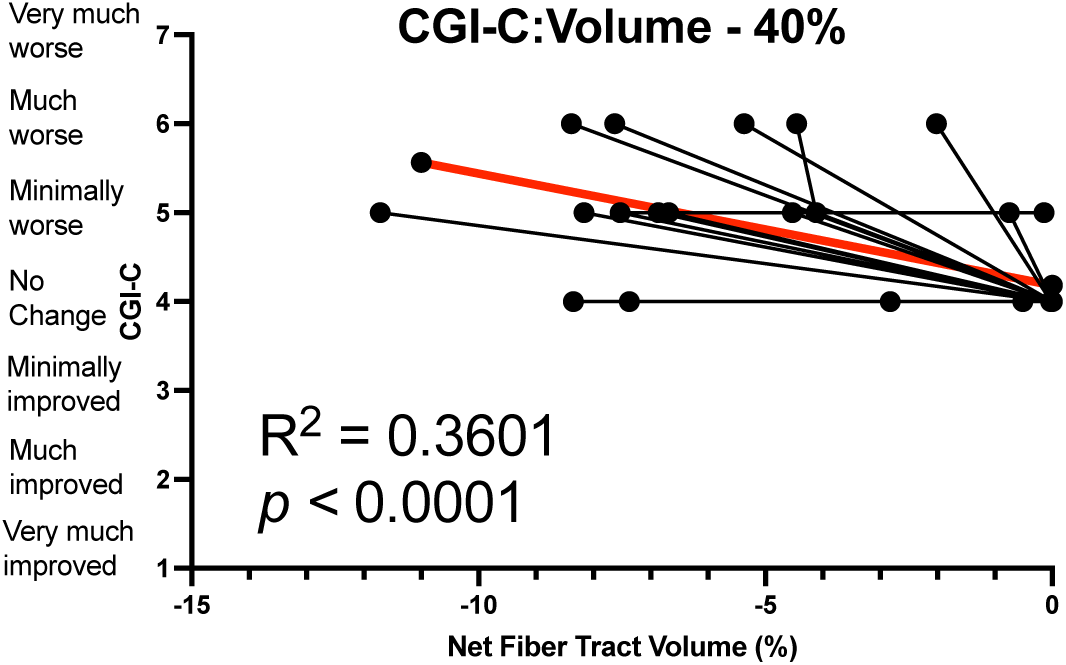
Differential Tractography correlations of net fiber tract volume with CGI-C change scores with GM1 patients at a 50% fractional anisotropy threshold.

**Table C1.**
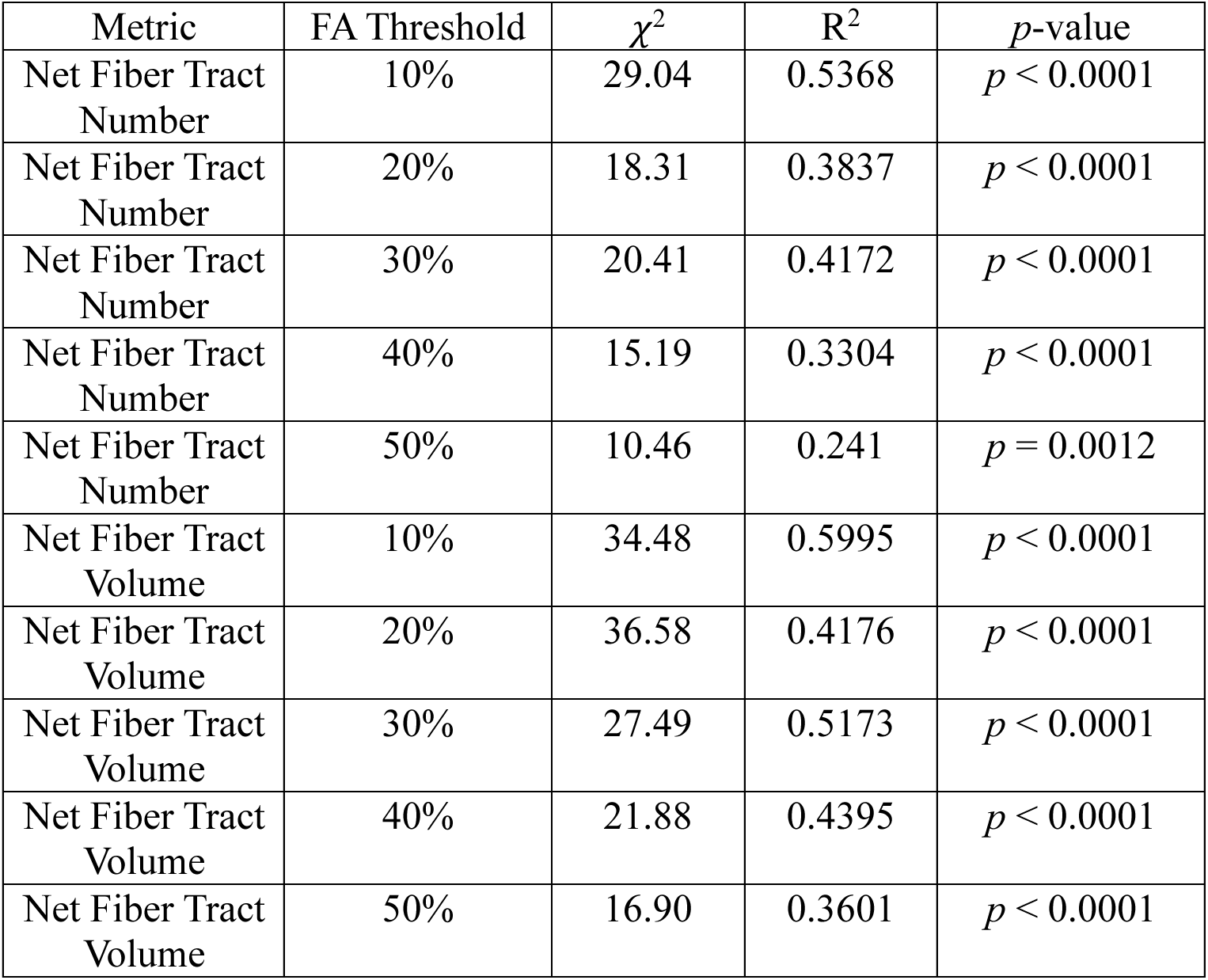
Correlations between net fiber tract number and net fiber tract volume with longitudinal CGI-C scores at varying fractional anisotropy thresholds.

